# Fundamental limits to the effectiveness of traveler screening with molecular tests

**DOI:** 10.1101/2024.07.11.24310291

**Authors:** Kate M. Bubar, Casey E. Middleton, Daniel B. Larremore, Katelyn M. Gostic

## Abstract

Despite the appeal of screening travelers during emerging infectious disease outbreaks, evidence shows that syndromic and questionnaire-based programs are largely ineffective in preventing or delaying the geographic spread of infection. Molecular tests offer high sensitivity and specificity, and can detect infections earlier than symptom screening, suggesting potential for improved outcomes, yet molecular tests were used to screen travelers for COVID-19 with mixed success. To investigate why screening for COVID-19 was not more successful, and to quantify the limits of screening for other pathogens of concern, we developed a probabilistic model that incorporates within-host viral kinetics. We then evaluated the potential effectiveness of screening travelers with molecular tests for influenza A, SARS-CoV-1, SARS-CoV-2, and Ebola virus. Even under highly optimistic assumptions about behavior and test characteristics, we find screening effectiveness is always limited because the infections with the highest transmission potential are undetectable at the time of travel, an idea we term the fundamental limit of traveler screening. We also demonstrate how estimates of ascertainment are a misleading substitute for screening effectiveness because they overestimate reductions in transmission at the destination. Understanding these limitations can guide the deployment of future traveler screening programs and inform strategies to improve outbreak prevention and control.

## Introduction

Air travel is a major driver of the geographic spread of emerging infectious diseases, directly linked to the international spread of SARS in 2003, influenza A/H1N1 in 2009, and SARS-CoV-2 in 2020 [1–3] as well as the importation of cases of influenza A/H7N9, MERS-CoV, Ebola, Lassa fever and Chikungunya [1, 4–6]. Although screening travelers for symptoms of infection may therefore seem like an intuitive countermeasure, scenario modeling [1, 3, 4, 7, 8] and overwhelming empirical evidence [9–13] show that syndromic and questionnaire-based screening programs are ineffective. For example, one modeling study found that even with a theoretical symptom-based test with perfect sensitivity, fewer than 3%, 9%, 10% and 35% of infected travelers would be detected for Ebola, SARS-CoV-2, SARS-CoV-1 and influenza, respectively [14].

An infected traveler may be missed by screening for two reasons. First, their infection may be undetectable at the time they are screened. For instance, syndromic screening will fail to identify asymptomatic and pre-symptomatic travelers. Second, their infection may be detectable in principle, yet missed because of imperfect test sensitivity. For instance, a thermometer may fail to detect a fever, or a person’s symptoms may differ from those surveyed. Novel rapid molecular tests appear to address both of these issues, offering high sensitivity and specificity over a long detectable window with rapid turnaround. These observations lead us to ask whether there are scenarios in which screening travelers for an infectious disease with a state-of-the-art molecular test could be effective in preventing or delaying an outbreak at the travelers’ destination.

Unfortunately, empirical evidence is limited. While molecular tests were used for traveler screening during the COVID-19 pandemic [15, 16], the jurisdictions that most successfully prevented or delayed transmission of SARS-CoV-2 such as New Zealand, Australia, Hong Kong, and Taiwan also had strict border controls, post-arrival quarantine measures, widespread testing, or contact tracing. As a result, it is unclear what role molecular testing of travelers per se played in practice, and what little evidence we do have (reviewed in [17]) covers only the number of individuals screening positive, but not programs’ effectiveness or impact on delaying transmission.

In place of empirical data, modeling studies offer various estimates for SARS-CoV-2 in particular. For example, a PCR test within the 24 hours before departure is predicted to reduce the number of infectious or pre-infectious travelers by 31% [18], while a PCR test within 3 days of departure is predicted to reduce the cumulative number of infectious days over the travel period by 36%, and would identify 88% of actively infectious travellers on the day of the flight [19]. Screening at airports has been projected to reduce post-arrival transmission risk by 37-47% for rapid diagnostic tests (RDTs) [20] or 28-50% for PCR or RDTs [21].

Here, in order to understand why molecular tests do not perform better in traveler screening, despite their high sensitivity, we introduce a probabilistic model that incorporates within-host viral kinetics to evaluate the effectiveness of screening. To explore how effective traveler screening could be in different settings and against different pathogens, we apply the model to analyze four example pathogens: influenza A, SARS-CoV-1, SARS-CoV-2, and Ebola virus. This modeling framework incorporates variation in individuals’ post-travel transmission potential based on variability in viral load trajectories and differences in when individuals travel during their infection. It is generalizable and could be adapted for other pathogens or other testing settings like pre-event screening.

## Results

### Model for traveler screening

From a public health perspective, the most important infections for a traveler screening program to catch are those most likely to infect others during or after travel, and the least important are those with little to no remaining infectiousness. To incorporate this concept into a mathematical model, we quantified an individual’s post-travel transmission potential using a simple within-host viral kinetics model (Fig. 1). We define individual *i*’s post-travel transmission potential, *R_i_*(*t^∗^*), as the expected number of secondary infections generated after traveling at time *t^∗^*. This approach accounts for variation in individual reproductive numbers and infection age at time of travel.

**Figure 1:**
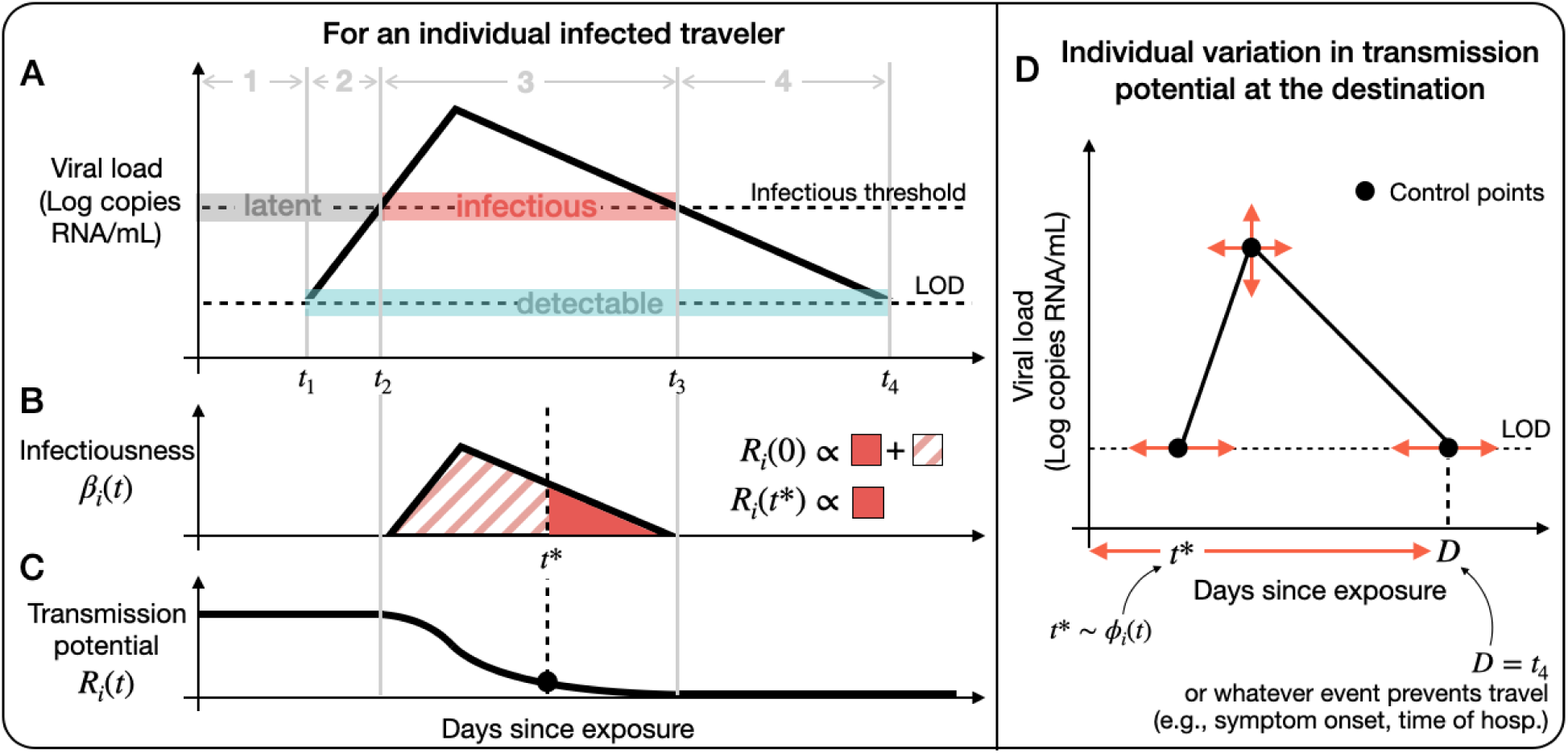
Model diagram. An example (**A**) viral load, (**B**) infectiousness *β_i_*(*t*), and (**C**) transmission potential *R_i_*(*t*) for an individual infected traveler *i*, with travel time *t^∗^* and post-travel transmission potential *R_i_*(*t^∗^*). There are four possible statuses for infected travelers: (1) not yet detectable or infectious, (2) detectable and not yet infectious, (3) detectable and infectious, (4) detectable and no longer infectious. (**D**) Factors that contribute to variation in *R_i_*(*t^∗^*): Stochastic realizations of viral load control points (first and last time detectable, peak viral load), when people may travel [0*, D*], and the simulated travel time *t^∗^* drawn from *ϕ_i_*(*t*), the infection age distribution among infected travelers.

We considered two different approaches to quantify traveler screening effectiveness. First, we considered how many additional infected travel attempts could be tolerated before causing an outbreak with high probability in screening vs no-screening scenarios (Δ*N*). To calculate Δ*N*, we used theory from stochastic processes about the long-term probability of extinction to compute the number of infected travelers required to cause an outbreak with probability *p* = 0.9. Second, we considered how much longer it takes for an outbreak of size *X* to occur at the destination in screening vs no-screening scenarios (Δ*t*). To calculate Δ*t*, we assumed that the number of infected travelers arriving at the airport followed a Poisson process with mean *λ* infected travelers per day. We simulated transmission chains initialized by infected travelers at the destination until *X* infections had occurred.

Throughout this work, we intentionally made optimistic assumptions about test performance, assuming instantaneous tests results, perfect compliance, and a limit of detection (LOD) equal to that of RT-PCR (hereafter PCR), the gold standard LOD currently achievable for the diseases in this study. We assumed 100% sensitivity above the limit of detection. For currently available technology, these assumptions are unrealistic because there is a tradeoff between sensitivity and turnaround time: PCR tests do not give instantaneous results and rapid tests are less sensitive [22]. However, these optimistic assumptions allow us to characterize the best-case scenario, and thus the *potential* effectiveness of screening programs.

In our comparisons of screening vs no-screening scenarios, we made two additional modeling assumptions. First, we considered screening only at points of exit, rather than paired screening at points of exit and entry. This is because prior work has found little marginal benefit for an additional test at points of entry, provided that the screening method is highly sensitive [1, 7]. Second, we assumed that the outbreak at the departure location is in a phase of exponential growth, an assumption relevant to screening-based containment scenarios, and one which affects the demographic distribution of infection ages among those attempting travel.

### Screening effectiveness to delay transmission

We simulated 5,000 infected travelers for four example pathogens: SARS-CoV-1, SARS-CoV-2, influenza A, and Ebola. We chose these pathogens because their diverse natural histories probe distinct areas of the parameter space of our model, and because traveler screening programs have been implemented for all of them. Then, we ran 20,000 simulations of the traveling process, sampling new infected travelers arriving at the airport from the 5,000 travelers until an outbreak was likely triggered (Fig. 2A, B). To calculate the time until an outbreak of size *X* occurs at the destination, we considered a plausible scenario for each pathogen. For SARS-CoV-1, SARS-CoV-2, and influenza A, *X* = 100 infections and *λ* = 1 infected traveler attempting travel per day on average. For Ebola, *X* = 1 infection and *λ* = 2 per month. We considered a different scenario for Ebola because, due to the severity of symptoms, any local transmission would be of concern, and sustained local transmission is relatively rare so *X* = 100 is less appropriate. We also expect the arrival rate of infected travelers to be less frequent because Ebola’s generation interval is much longer than respiratory pathogens and non-pharmaceutical interventions can usually bring Ebola transmission under control, or nearly so.

**Figure 2:**
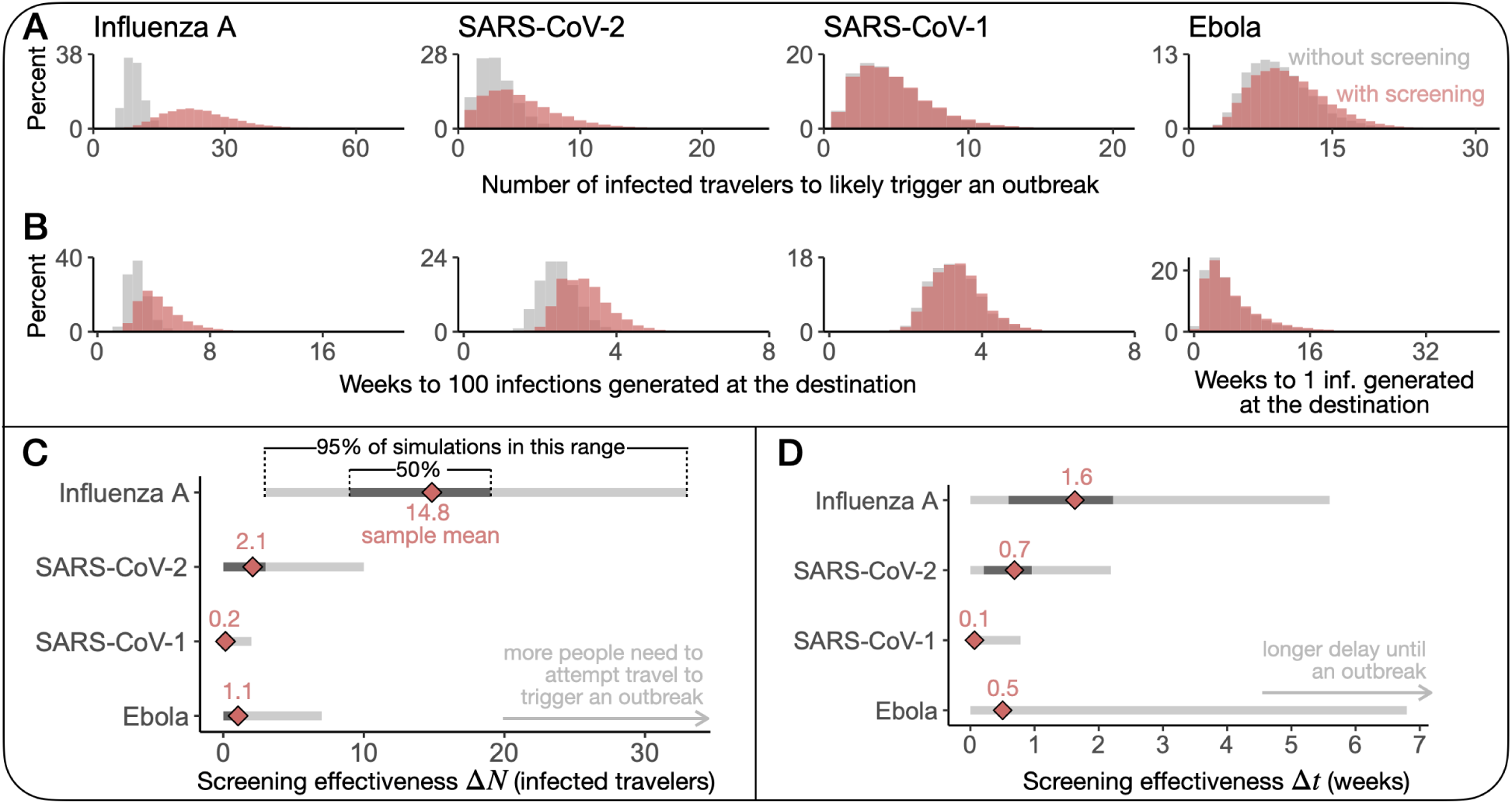
Screening effectiveness to delay transmission is limited and highly variable. Histograms of (**A**) the number of infected travelers to likely trigger an outbreak with (pink) and without screening (gray) and (**B**) the time to *X* infections generated at the destination with (pink) and without screening (gray) from 20,000 Monte Carlo simulations. *X* = 100*, λ* = 1 per day for SARS-CoV-1, SARS-CoV-2, and influenza A. *X* = 1*, λ* = 2 per month for Ebola. (**C, D**) Distributions of Δ*N* and Δ*t* from 20,000 Monte Carlo simulations (sample mean (pink diamond), IQR (dark gray) and 95% percentile range (light gray)).

Of the four pathogens we considered, traveler screening is most effective for influenza A. With screening in place, it takes an average of 15 more infected individuals to attempt travel to trigger an outbreak in comparison to no screening program (Fig. 2C). In units of time, screening delayed an influenza A outbreak at the destination by 11.1 days on average (Fig. 2D). However, there is considerable variation in both of these outcomes, so it is important to consider their distributions when interpreting these results. One way to understand this variation is to consider the range of the outcomes, such as the range of the central 50% of simulations (i.e., the interquartile range; Fig. 2C, D). Another way to understand this variation is to compute the probability that screening delayed an outbreak by at least *x* infected travelers or *x* days. For example, screening delayed an outbreak by at least one week in 57.6% of simulations for influenza A. See Supp. Table S1 for more values of *x*.

Traveler screening was less effective for the other three pathogens. The average Δ*N* was 0.2, 1.1, and 2.1 infected travel attempts for SARS-CoV-1, Ebola, and SARS-CoV-2, respectively. Screening delayed outbreaks by 0.4, 3.5, and 4.8 days on average for SARS-CoV-1, Ebola, and SARS-CoV-2, respectively (Fig. 2D). Once again, it is important to consider the variation in these outcomes. For example, although the average value of Δ*t* for Ebola is around half a week, the Δ*t* distribution is highly skewed—in over 50% of simulations, there is no delay at all. Screening delayed an outbreak by at least a week in 1.4%, 9.3%, and 23.4% of simulations for SARS-CoV-1, Ebola, and SARS-CoV-2, respectively.

### Fundamental limit of traveler screening

Ideally traveler screening would prevent an outbreak at the destination, or delay transmission long enough to enable some public health activation. However, even for influenza A, which had the most effective traveler screening programs on average, individual trial results varied greatly, and sometimes screening did not delay transmission at all (Fig. 2C, D). Why is screening not more reliably effective, especially under highly optimistic assumptions about detectability and test sensitivity?

To answer this question, we observe that, for any test or pathogen, there always exists a gap between when someone is infected and first detectable. This implies that there is a window of time when an infected individual may travel and is undetectable at the airport. Crucially, travelers who are missed by screening during this window have all of their transmission potential remaining (Fig. 3A). Thus, the travelers with the most transmission potential are impossible to catch.

**Figure 3:**
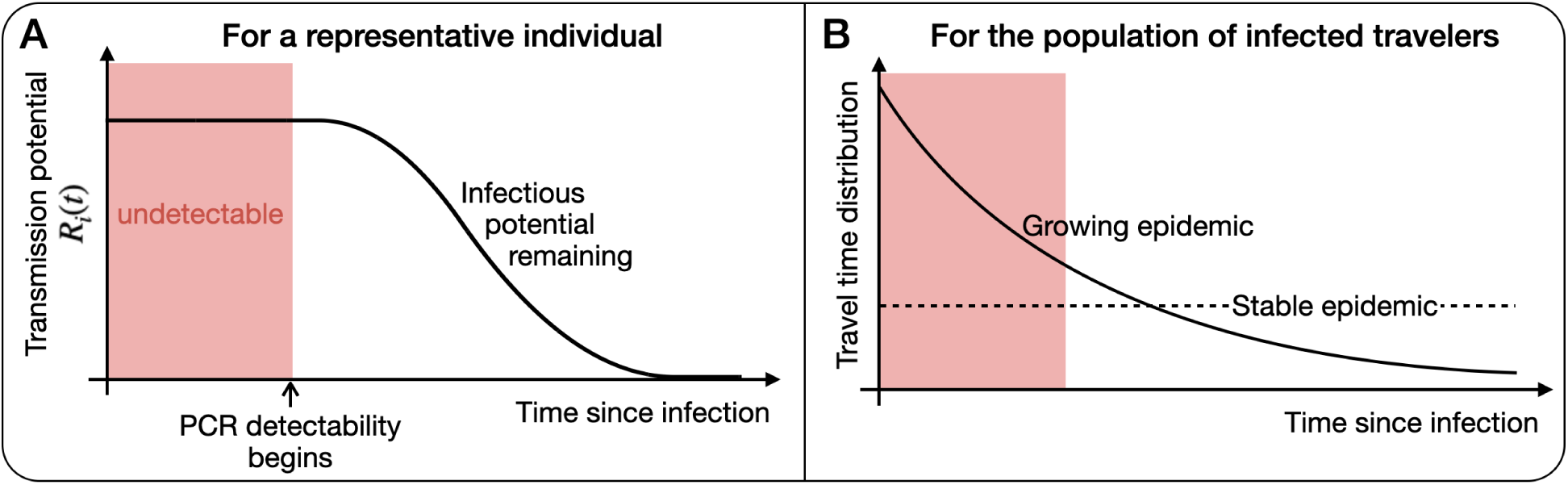
The effectiveness of screening travelers is fundamentally limited by the gap between infection and detectability. **(A)** Individuals are undetectable by molecular testing when their transmission potential is highest, fundamentally limiting the effectiveness of traveler screening because infected people may travel during this window. **(B)** A growing epidemic exacerbates this fundamental limit because the infection age distribution among infected travelers is positively skewed in comparison to a stable epidemic.

To quantify how this window of time limits traveler screening effectiveness, we calculated the expected proportion of transmission potential that is detectable by screening,

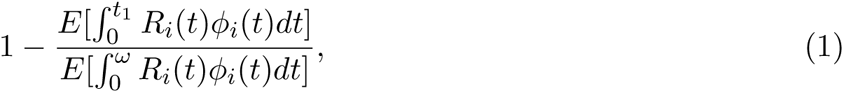

for all individuals *i* with transmission potential. Using the notation from Fig. 1, *t*_1_ is the time *i* is first detectable, and *ω* is the time *i* is either no longer infectious or no longer able to travel (*ω* = min(*t*_3_*, D*)). The infection age distribution among infected travelers, *ϕ_i_*(*t*), is a mixture of the infection age distribution and the propensity to travel at a particular age.

We found that, during a growing epidemic, only 2.8%, 9.7%, 40.2% and 59.8% of transmission potential is expected to be detectable by traveler screening via molecular test for SARS-CoV-1, Ebola, SARS-CoV-2, and influenza A, respectively. Our estimate for SARS-CoV-2 is comparable to other modeling studies that found testing reduced post-arrival transmission risk by 29–53% [20, 21].

Expression 1 represents a fundamental limit to the effectiveness of traveler screening. Because of the gap between infection and detectability, the fraction in Expression 1 is always positive. Consequently, traveler screening alone can never eliminate the risk of local transmission at the travel destination. Furthermore, during a growing epidemic, most of the infected traveling population would travel during this early window of undetectability coincides (Fig. 3B), exacerbating the consequence of this fundamental limit.

### Ascertainment overestimates transmission reduction

Although difficult to measure in practice, a common approach in modeling studies to evaluate the effectiveness of a screening program is to estimate ascertainment, the percentage of infections detected out of all the infections screened. Out of the 5,000 simulated travelers, we found that ascertainment is extremely low for SARS-CoV-1 and Ebola (3.1% and 10.5%, respectively), and better but still imperfect for SARS-CoV-2 and influenza A (47.8% and 70.9%, respectively). Of note, ascertainment is unaffected by test turnaround time, allowing us to compare our estimate of SARS-CoV-2 ascertainment to an empirical estimate from testing at U.S. airports during 2022 (52%) [23], a difference of just 4.2 percentage points.

The ascertainment rate is a misleading substitute for screening effectiveness because it overestimates reductions in transmission at the destination. This is because the typical undetected traveler has a greater post-travel transmission potential than the typical detected traveler, a consequence of the fundamental limit of traveler screening. We found that the average *R_i_*(*t^∗^*) among undetected travelers is 2.6, 1.8, 2.5, and 1.2 for SARS-CoV-1, Ebola, SARS-CoV-2, and influenza A, respectively, while for detectable travelers, it is 2.8, 1.7, 1.8, and 0.8. This pattern occurs because many of the detected infected travelers are near the end of their course of infection and have little to no transmission potential (Supp. Fig. S1). Moreover, for all four pathogens, the percent of post-travel transmission potential that is detectable by screening is always less than the corresponding ascertainment rate.

### Sensitivity Analysis

The effectiveness of traveler screening is always limited by the amount of time people are infected before they are detectable. How variable is such imperfect screening effectiveness across different epidemic scenarios, test characteristics, infectiousness profiles, and traveling behaviors?

Across various epidemic scenarios (*λ* = *{*1 per day, 1 per week, 2 per month*}*, *X* = *{*1, 10, 100*}* infections), we intuitively found that screening delays outbreaks for longer when infected people travel less frequently or when the outbreak threshold is larger (Supp. Figs. S2, S3, S4, and S5). However, even in the best case scenario (*λ* = 2 per month, *X* = 100), Δ*t* remains highly variable. For example, in this scenario, the average delay to 100 influenza A infections is 95.8 days, yet Δ*t* is less than one week in 38.9% of simulations (Supp. Fig. S2). We found negligible differences in Δ*N* and Δ*t* when modeling the rate at which infected people attempt travel as either a homogeneous or non-homogeneous Poisson process as in [7].

Infectious thresholds are estimated in the literature for SARS-CoV-2 and influenza A (Table S2) but not for SARS-CoV-1 or Ebola. For these pathogens, we chose infectious thresholds so the distributions of *R_i_*(0) are similar to the gamma distribution with mean *R*_0_ and dispersion parameter *k* [24] and ran sensitivity analyses with thresholds 10*×* larger and 10*×* smaller. A lower infectious threshold decreases the amount of time it takes to generate *X* infections at the destination because more infected travelers have post-travel transmission potential (Supp. Fig. S6). However, the change in the average Δ*N* and Δ*t* due to different infectious thresholds was at most 1 person or 2.4 days (Supp. Figs. S7, S8, S9, and S10).

Finally, we assumed the probability an individual travels is uniform from infection to viral clearance for SARS-CoV-2 and influenza A. For SARS-CoV-1 and Ebola, we assumed symptoms prevent travel, limiting travel from infection to the time of hospitalization. If symptom severity did not impede travel for these pathogens, screening would be more effective because travelers are more likely to be detectable. Under these assumptions, the mean Δ*t* increased from 3.5 to 7.9 days for Ebola, (Supp. Fig. S11) and from 0.5 to 3.8 days for SARS-CoV-1 (Supp. Fig. S12).

## Discussion

This study modeled the potential effectiveness of traveler screening programs with highly sensitive molecular diagnostics to delay transmission at the destination. Overall, we found that screening effectiveness is generally quite limited, or at best, highly variable. Of the four pathogens we considered, traveler screening was most effective for influenza A, but even under our optimistic assumptions about test performance, over 40% of post-travel transmission potential is *not* preventable by screening. This limitation is exemplified by what we refer to as the fundamental limit of traveler screening. The idea is simple: the effectiveness of traveler screening programs will always be limited because, for every diagnostic test and pathogen, the newest infections with the most remaining transmission potential are impossible to catch. Even with state-of-the-art tests where people are detectable before they are infectious, there is a window of time after the infection event when individuals are not yet detectable and may travel, and their infectious period will not begin until they are at the destination.

The consequences of this fundamental limit are exacerbated during a growing epidemic, precisely when traveler screening programs would likely be implemented, because infections are more likely to be recent. This fact reinforces the idea that any post-screening countermeasures such as arrival quarantines should be sized and scoped for the full duration of when infections are likely undetectable [21], or in the case of syndromic screening, the full incubation period [14].

The fundamental limit can help us understand when traveler screening programs are more likely to be effective. The best case scenario for screening would involve a pathogen such that people are detectable very quickly after infected (ideally within hours [14]) or, if individuals are undetectable for a long period of time, they have low post-travel transmission potential (i.e., *t*_1_ and *R_i_*(*t*) are negatively correlated). Additionally, screening is more likely to be effective if transmission is highly dispersed simply because most individuals who are missed by screening are unlikely to infect others [24]. Importantly, this notion of controllability differs from that of Fraser et. al. [25] and Middleton and Larremore [26], because effective control of community transmission requires that detectability precede infectiousness. While this requirement is necessary for effective traveler screening, it is not sufficient. One also needs there to be no undetectable window after exposure.

While molecular tests are typically more sensitive than syndromic screening, they are not always superior for traveler screening. For example, we found traveler screening via molecular tests is extremely ineffective for SARS-CoV-1, even though it is considered a controllable pathogen [25]. This discrepancy is due to symptom onset occurring before infectiousness and typically before detectability by PCR. In the initial days post symptom-onset, 50-80% of infections are negative by PCR with nasopharyngeal aspirate samples [27–32], possibly because viral replication starts in the lower respiratory tract [27]. Thus, effective screening depends on the natural history of the disease.

This work assumes the goal of traveler screening is to delay or prevent local transmission at the destination, yet screening may also be used for general surveillance, sequencing, and public awareness of an ongoing outbreak. For example, in Venezuela in 2021, the introduction of the SARS-CoV-2 Omicron variant was rapidly detected in samples from airport screening [33]. Our work cautions that measurements of prevalence among travelers are likely to be poor, even with a state-of-the-art test, yet could be corrected using our model’s ascertainment estimates.

Our findings are subject to a number of limitations. First, the exact effectiveness of screening programs will depend on whether our model truly captures viral kinetics and infectiousness profiles. The data available to parameterize viral load trajectories vary widely in quality and quantity. For example, the distributions of control points for the viral load function are well characterized for SARS-CoV-2, but meager or nonexistent for other pathogens. In these cases, we used the best estimates available or extrapolated plausible ranges from other available information like the percent of PCR positivity upon hospitalization (Table S2). Additionally, while the use of log viral load as a proxy for infectiousness is supported in the literature (SARS-CoV-1 [25, 34], SARS-CoV-2 [35, 36], Ebola [37], influenza A [38]), other relationships between viral load and infectiousness [22, 39], or other proxies for infectiousness [40, 41], are possible.

Another limitation of this model is that the simplistic model of *β_i_*(*t*) assumes that all variation in individuals’ transmission potential is due to their viral load, and not differences in social contacts or behavior. This model also excludes other modes of transmission like post-mortem transmission that is known to be an important driver in the spread of Ebola. Improved characterization of individual viral load trajectories, and how they relate to infectiousness, symptoms and behavior, would greatly improve the impact and value of this, and other [22, 26, 42], modeling studies. Moreover, while the simple triangular model of *β_i_*(*t*) is used in other modeling studies [26, 43], the modeling framework is flexible and more sophisticated functional forms of *β_i_*(*t*) could capture time-varying contact patterns or behavior.

We assumed tests have a highly sensitive PCR limit of detection, around 10^2.6^ copies of RNA/mL (Table S2). One way to improve screening effectiveness would be to design tests with a lower limit of detection so individuals are detectable earlier. Lower PCR limits of detection are physically possible, for example by using more sensitive PCR enzymes or optimizing the PCR conditions, but reliability at lower thresholds is challenging. In practice, any operational delays in sample-to-answer times would substantially lower screening effectiveness, so our results, which assumed instantaneous results, represent an upper bound on the potential effectiveness of traveler screening.

More broadly, our work is situated within a family of research that uses mathematical modeling to evaluate different testing scenarios. Other areas that our modeling framework could easily extend to include pre-event testing [20, 42], screening in combination with other interventions such as quarantine and/or contact tracing [3, 18, 19, 21], and combining multiple screening methods such as fever screening and health questionnaires with molecular testing [1, 4].

Traveler screening programs are typically expensive and resource intensive to implement. Our results suggest that, while traveler screening may delay an outbreak at the destination, combining traveler screening with other interventions is necessary to more consistently delay, or ideally prevent, an outbreak post-travel. Unfortunately, screening travelers with more sophisticated rapid molecular diagnostics will not be as effective as hoped at delaying transmission because the travelers with the highest transmission potential are likely impossible to detect.

## Methods

### Approximation of *R_i_*(*t*)

Let *β_i_*(*t*) be the infectiousness of a single individual *i* at time *t* during their course of infection. We assume that *β_i_*(*t*) reflects infectiousness and an average over typical behavior in the absence of interventions. Mathematically,

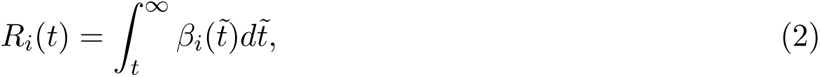

where *R_i_*(*t*) is the expected number of secondary infections *i* will generate after time *t*. Here, *R_i_*(0) is *i*’s individual reproductive number and the population-level basic reproductive number *R*_0_ = *E*[*R_i_*(0)]. The individual reproductive number is also commonly referred to as *ν* [24] or simply *R_i_*. *R_i_*(*t*) is a monotonically decreasing function, which is biologically realistic: the number of expected secondary transmission events ahead in time decreases as an individual’s infection progresses (Fig. 1C).

To compute *R_i_*(*t*), we approximate *β_i_*(*t*) using a simple within-host viral kinetics model with an infectious threshold as a proxy for infectiousness over time (Fig. 1A, 1B). The within-host model assumes there is a period after infection where the virus is undetectable, and then a proliferation phase of exponential growth followed by a clearance phase of exponential decay. This type of log-linear proliferation and clearance model, sometimes referred to as a hinge or tent function, is commonly used to describe the proliferation and clearance phase of viral infection [22, 26, 42, 44, 45]. We assume there is an infectious threshold such that infectiousness is proportional to log viral loads above this threshold.

For SARS-CoV-2 and influenza A, we found estimates for the infectious threshold in the literature (Table S2). For SARS-CoV-1 and Ebola, we estimated infectious thresholds to result in distributions of *R_i_*(0) similar to the gamma distribution with mean *R*_0_ and dispersion parameter *k*, a typical choice for the distribution of individual reproductive numbers [24]. Fitting the infectious threshold directly to the gamma distribution would assume that all the variation in individual reproductive numbers is due to differences in viral loads. However, we know that other factors contribute to differences in individual reproductive numbers so we would not expect the distribution of *R_i_*(0) from the viral load model to identically match the distribution of *R_i_*(0) fit to contact tracing data. We checked how sensitive our results were to the infectious threshold value in the sensitivity analyses (Supp. Fig. S6, S7, S8, S9, S10).

### Simulations

Each simulated infected traveller is assigned a time they are first and last detectable by a molecular test with a PCR limit of detection, a time and magnitude of peak viral load, and a time of hospitalization (for SARS-CoV-1 and Ebola) sampled from the distributions in Table S2. Some of these distributions are hard to estimate in practice so the distributions used were optimistic estimates, in terms of potential screening effectiveness, informed by existing literature when well-characterized distributions were not available (Described further in the Supp. Materials). Individuals’ travel times *t^∗^* are sampled from the infected travelers’ infection age distribution *ϕ*(*t*) using the inverse CDF method. With these parameters, we compute individuals’ *R_i_*(*t^∗^*) and screening result at the time of travel.

For each individual, we simulate their contribution to infection at the destination using a branching process in which the offspring distribution of the first generation is a Poisson distribution with *λ* = *R_i_*(*t^∗^*), and for subsequent generations, a Negative binomial distribution with mean *R*_0_ and a disease-specific dispersion parameter *k* [24]. If *i* is not detected, the simulated branching processes are identical with and without screening. This approach of comparing counterfactual scenarios ensures our results reflect the impact of screening alone and not the stochasticity of transmission.

### Number required to likely trigger an outbreak

Following Clifford et. al. [7], we can estimate the long-term probability of disease extinction *s*_0_, for a negative binomial offspring distribution with mean *R*_0_ and dispersion parameter *k*, from the implicit equation

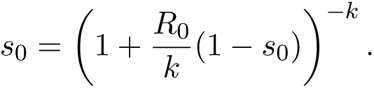

Let *s_i_* be the long-term probability of disease extinction in a population where the first generation of infections is caused by infected traveler *i* with transmission potential *R_i_*(*t^∗^*), with offspring distribution Poisson(*R_i_*(*t^∗^*)), and each subsequent generation follows NegBinom(*R*_0_*, k*). Then,

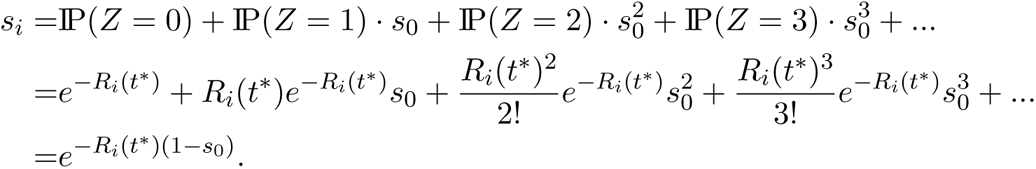

As in [7], the probability that infected traveler *i* causes an outbreak at the destination is *q_i_*= 1 *−s_i_*.

To calculate *N*, the number of infected travelers required to trigger an outbreak, we know that the first *N −* 1 travelers did not cause an outbreak. If *q_i_* = *q* for all infected travelers, *X ∼* geometric(*q*) so

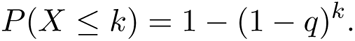

However, since *q_i_*is dependent on an individual’s *R_i_*(*t^∗^*), *q_i_* is a random variable so

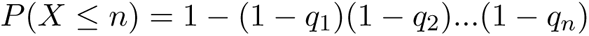

for *n* infected travelers. For each run of our model simulation, we can use this equation to compute the number of infected travelers *N* required to cause an outbreak with probability *p*. For our analyses, we set *p* = 0.9.

### Time to *X* infections generated at the destination

To compute this outcome via simulation, we first generate an arrival time for an infected traveler and simulate any transmission chains they generated using the distributions described above. We store the first *X* subsequent cases and the timing of infection using the pathogen-specific generation interval. Then, we generate the arrival time of the next infected traveler. If this infected traveler arrived before the last stored case, or the number of cases at the destination is less than *X*, we repeat these steps until the requirements have been met. The output, time to *X* infections generated at the destination is the time of the *X^th^* infection.

### Infected travelers’ infection age distribution

We assume traveler screening programs would be implemented at the beginning of an emerging infectious disease outbreak when infections are growing exponentially. So, as previously described in [1], the probability that an infected traveler has infection age *t* at the time of travel is

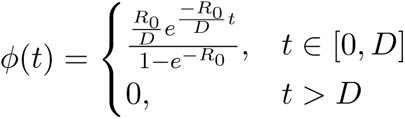

where *D* is the duration of infection in which an infected individual is assumed to travel. If the disease does not prevent someone from traveling, *D* is the time from infection to viral clearance. If symptoms prevent an individual from traveling, we assume *D* is the average time from infection to hospitalization.

The corresponding CDF is the probability that an infected traveler was infected less than or equal to *t* days before travel,

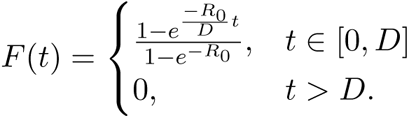

## Data Availability

All code used in these analyses can be found online.

https://github.com/kbubar/travelerscreening

## Acknowledgements

The authors thank Stephen Kissler and Ellen DeGennaro for their feedback.

## Code and data availability

All code used in these analyses can be found at https://github.com/kbubar/travelerscreening.

## Supplemental Materials

### Viral load parameterization

Table S2 contains all of the parameter values and distributions used to simulate infected travelers for each pathogen. For each parameter that is treated as a random variable, we used the reported distribution when reported in the literature. If we could not find a reported distribution, we used a truncated normal when a mean and standard deviation were reported, truncated either at the lowest and highest reported values or within a reasonable range that captured the vast majority of measurements. If mean and SD were not reported, then we used a uniform distribution within a reasonable range that captured the vast majority of measurements. We used the serial interval as an approximation for the generation interval when generation interval estimates were not available. Examples of 100 simulated viral load trajectories for these four pathogens are shown in Supp. Fig. S13. We chose the lowest reported PCR limit of detection, since this corresponds to a best case scenario for testing. Below we elaborate on specific assumptions and rationale for each pathogen.

### SARS-CoV-1

We assumed that all the cases are hospitalized. This is appropriate because hospitalization rates for symptomatic SARS-CoV-1 were high and, while estimates of asymptomatic infections vary from 0.1% [46] to 13% [47], there is no known transmission from asymptomatic patients so we do not consider them in our analyses [47]. We chose the distribution uniform(0,9) days post symptom onset as an optimistic guess for the time first detectable by PCR. This range was chosen based off data that reported 50-80% did not test positive via PCR in initial days post symptom onset [27–32], *>*50% were positive by day 6-7 [28, 29], and *>*95% are PCR positive by day 10 [32]. Although not necessary for the model, we also parameterized individuals’ time of symptom onset since other parameters were measured in units of the time since symptom onset.

### SARS-CoV-2

We parameterized the model for the ancestral strain of SARS-CoV-2. Many of the parameters needed for our model are well characterized by Kissler et. al. [42]. We did not distinguish between symptomatic and asymptomatic cases, nor did we include hospitalization.

### Influenza A

The majority of references reported data from influenza H1N1 subtype, a few from influenza H3N2, and some simply referenced influenza A without specifying the subtype. We did not distinguish between symptomatic and asymptomatic cases, nor did we include hospitalization.

### Ebola

For Ebola, higher viral load is correlated with mortality [48]. We parameterized the model for non-fatal cases assuming they would be more likely to travel and assumed all such cases are hospitalized. Although not necessary for the model, we parameterized individuals’ time of dry and wet symptom onset since other parameters were measured in units of the time since symptom onset. Note that the model parameters are not as well characterized for Ebola as other pathogens, possibly because the incubation and infectious periods are highly variable [49].

Our model of infectiousness implicitly assumes that we are only considering direct transmission via fomites, droplets, or aerosols [50], and not post-mortem transmission. Thus, we did not consider asymptomatic cases because they would not have transmission potential, and asymptomatic cases are rare [51].

We chose the distribution uniform(0, 3) days post onset of dry symptoms as an optimistic guess for the time first detectable by PCR. This range implies that individuals are sometimes detectable when they have dry symptoms, and everyone is detectable by the time their symptoms progress to wet symptoms. This was informed by the notion that there is no evidence that infected people are viremic before symptom onset, but some are PCR positive on the day of illness onset [52]. Additionally, most are detectable by the time they are hospitalized (87% [53]).

Note that viral load measurements are measured in mL of serum. This is appropriate for our model, since RDTs that could potentially be used for airport screening can collect a blood sample through a finger prick [54].

**Table S1:**
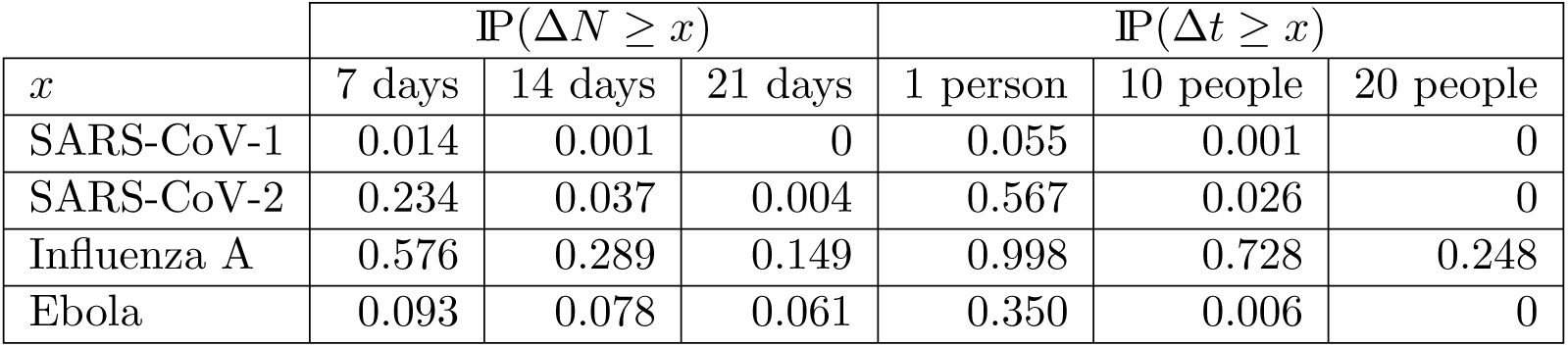
Examples of the complementary CDF IP(*X ≥ x*) for screening effectiveness Δ*N* and Δ*t*. For Δ*t*, we used the same scenarios as the main text where *X* = 100*, λ* = 1 for SARS-Cov-1, SARS-CoV-2 and influenza A, and *X* = 1*, λ* = 1*/*14 for Ebola.

| $x$ | $\mathbb{P}(\Delta N \geq x)$ | | | $\mathbb{P}(\Delta t \geq x)$ | | |
| --- | --- | --- | --- | --- | --- | --- |
|  | 7 days | 14 days | 21 days | 1 person | 10 people | 20 people |
| SARS-CoV-1 | 0.014 | 0.001 | 0 | 0.055 | 0.001 | 0 |
| SARS-CoV-2 | 0.234 | 0.037 | 0.004 | 0.567 | 0.026 | 0 |
| Influenza A | 0.576 | 0.289 | 0.149 | 0.998 | 0.728 | 0.248 |
| Ebola | 0.093 | 0.078 | 0.061 | 0.350 | 0.006 | 0 |

**Figure S1:**
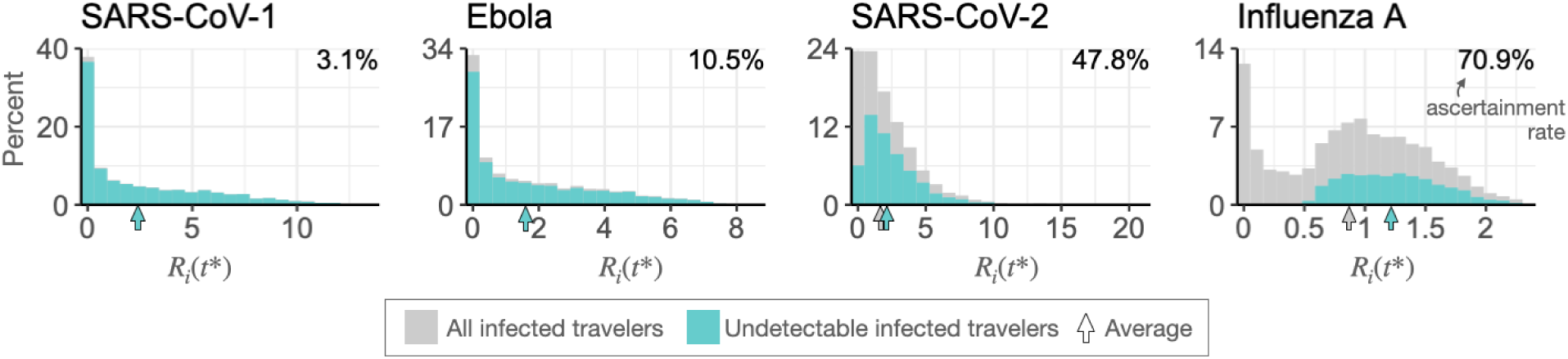
Traveler screening programs decrease the number of infected travelers reaching the destination, but the average imported case has more transmission potential than without screening. Histograms of simulated infected travelers’ transmission potential at the destination, *R_i_*(*t^∗^*), with all 5,000 travelers shown in gray and undetectable travelers in teal. Vertical arrows indicate the mean of each distribution. The means overlap for SARS-CoV-1 and Ebola. Ascertainment rates are reported in each upper right corner.

**Table S2:** Model parameters. Values or distributions used for each pathogen-specific parameter. SO stands for symptom onset.

| Pathogen | Parameter | Value/Distribution | Units | Source |
| --- | --- | --- | --- | --- |
| SARS-CoV-1 | time first detectable | Unif(0,9) | days from SO | [27–32] |
|  | time of peak VL | Unif(7, 14) | days from SO | [28, 32, 34, 55] |
|  | time last detectable | Unif(18, 28) | days from SO | [28, 29, 32] |
|  | PCR LOD | 2.6 | log10 copies/mL | [56] |
|  | infectious threshold | 6.5 | log10 copies/mL | See Methods |
|  | peak VL | Unif(5.8, 8.5) | log10 copies/mL | [32, 57–59] |
|  | time to symptoms | Unif(2,10) | days from infection | [60, 61] |
| | time to hospitalization | Trunc. normal ($\mu = 2.9$,<br>$\sigma = 2.6, a = -1, b = 9$) | days from SO | [60] |
| | $R_0$ | 2.55 | / | [24] |
| | $k$ | 0.21 | / | [24] |
|  | generation interval | 8.4 | days | [62] |
| SARS-CoV-2<br>(ancestral) | time first detectable | Unif(2.6, 3.8) | days since infection | [63] |
| | time of peak VL | $\Gamma(\text{shape}=2.3, \text{rate}=0.7)$ | days from first detect. | [42] |
| | time last detectable | $\Gamma(\text{shape}=2.4, \text{rate}=0.3)$ | days from peak | [42] |
|  | PCR LOD | 40 | CT | [42] |
|  | infectious threshold | 5 | log10 copies/mL | [42, 64] |
| | peak VL | Normal($\mu = 22.3, \sigma = 4.2$) | CT | [42] |
| | $R_0$ | 2.8 | / | [65] |
| | $k$ | 0.55 | / | [66] |
|  | generation interval | 5.9 | days | [67] |
| Influenza A | time first detectable | Unif(0.5, 1.5) | days from infection | [68, 69] |
|  | time of peak VL | Unif(1, 3) | days from first detect. | [68–70] |
|  | time last detectable | Unif(2, 3) | days from peak | [68–70] |
|  | PCR LOD | 2.95 | log10 copies/mL | [70] |
|  | infectious threshold | 4 | log10 copies/mL | [38] |
|  | peak VL | Unif(6, 8.5) | log10 copies/mL | [70] |
| | $R_0$ | 1.26 | / | [71] |
| | $k$ | 2.36 | / | [71] |
|  | generation interval | 2.6 | days | [72] |
| Ebola | time first detectable | Unif(0, 3) | days from SO | [73–75] |
|  | time of peak VL | Unif(3, 6) | days from SO | [76–79] |
| | time last detectable | Trunc. normal($\mu = 12.7$,<br>$\sigma = 3.8, a = 9, b = 16.5$) | days from SO | [76] |
|  | PCR LOD | 2.7 | log10 copies/mL | [78] |
|  | infectious threshold | 7 | log10 copies/mL | See Methods |
|  | peak VL | Unif(6.5, 9.2) | log10 copies/mL | [76, 77, 79] |
|  | time to dry symptoms | Unif(5, 13) | days from infection | [80, 81] |
|  | time to wet symptoms | Unif(3, 5) | days from SO | [74] |
| | time to hospitalization | Trunc. normal($\mu = 4.5$,<br>$\sigma = 2.5, a = 2, b = 7$) | days from SO | [76] |
| | $R_0$ | 1.8 | / | [82–84] |
| | $k$ | 0.18 | / | [84] |
|  | generation interval | 13 | days | [81] |

**Figure S2:**
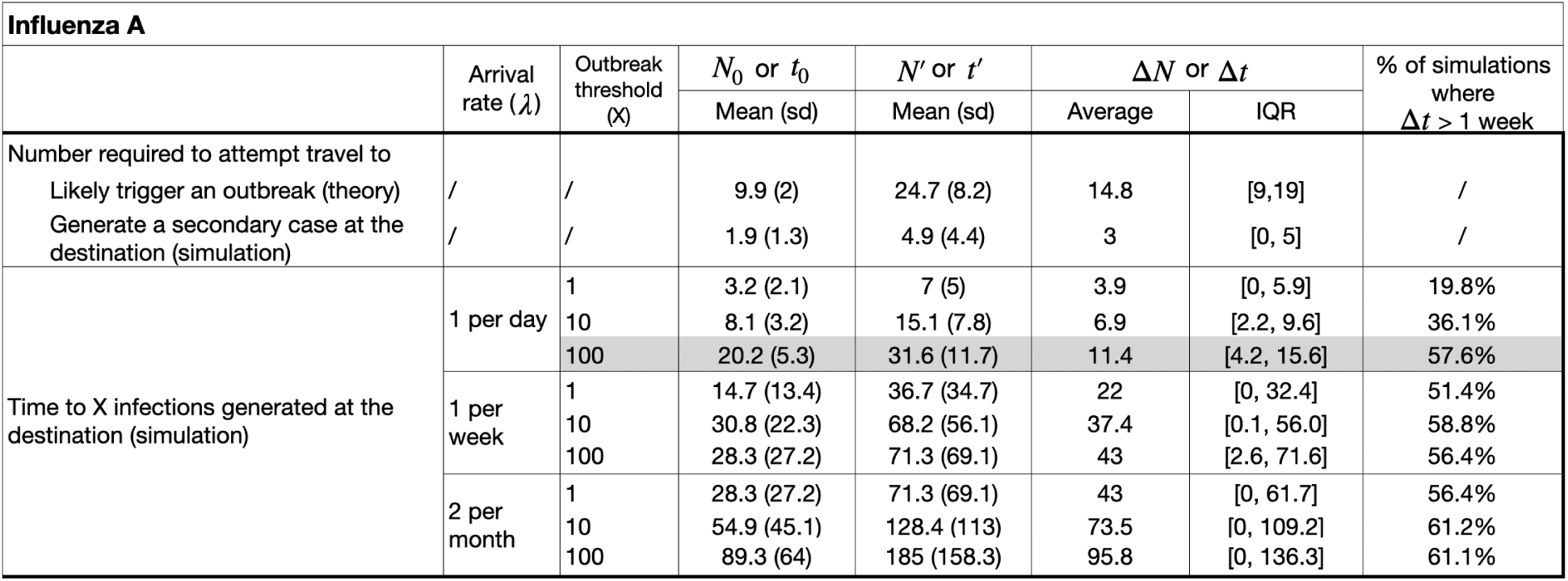
Screening effectiveness for influenza. **A.** The number of infected travelers required to attempt travel to likely generate an outbreak with and without screening (*N^′^* and *N*_0_, respectively), and the time to *X* infections generated at the destination for a range of *X* with and without screening (*t^′^* and *t*_0_, respectively), and screening effectiveness Δ*N* = *N^′^ − N*_0_ and Δ*t* = *t^′^ − t*_0_ for a range of scenarios. The gray row is the plausible example reported in the Main Text.

**Figure S3:**
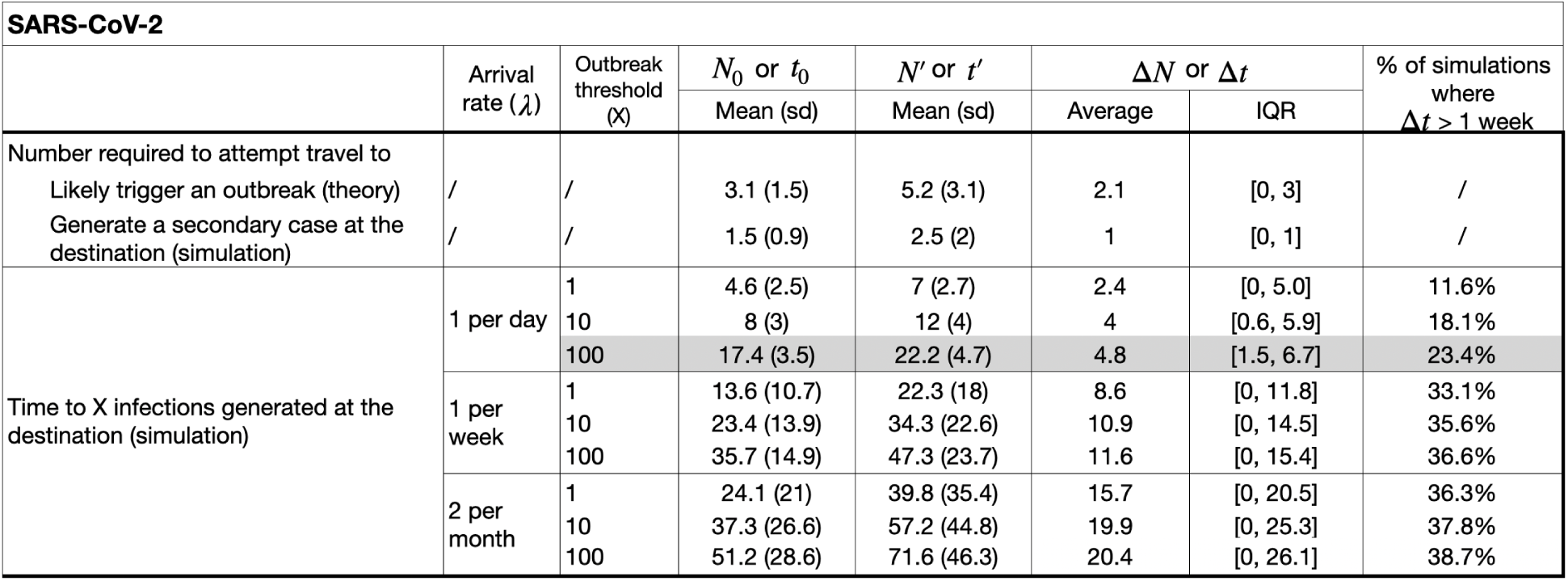
Screening effectiveness for SARS-CoV-2. The number of infected travelers required to attempt travel to likely generate an outbreak with and without screening (*N^′^* and *N*_0_, respectively), and the time to *X* infections generated at the destination for a range of *X* with and without screening (*t^′^* and *t*_0_, respectively), and screening effectiveness Δ*N* = *N^′^ − N*_0_ and Δ*t* = *t^′^ − t*_0_ for a range of scenarios. The gray row is the plausible example reported in the Main Text.

**Figure S4:**
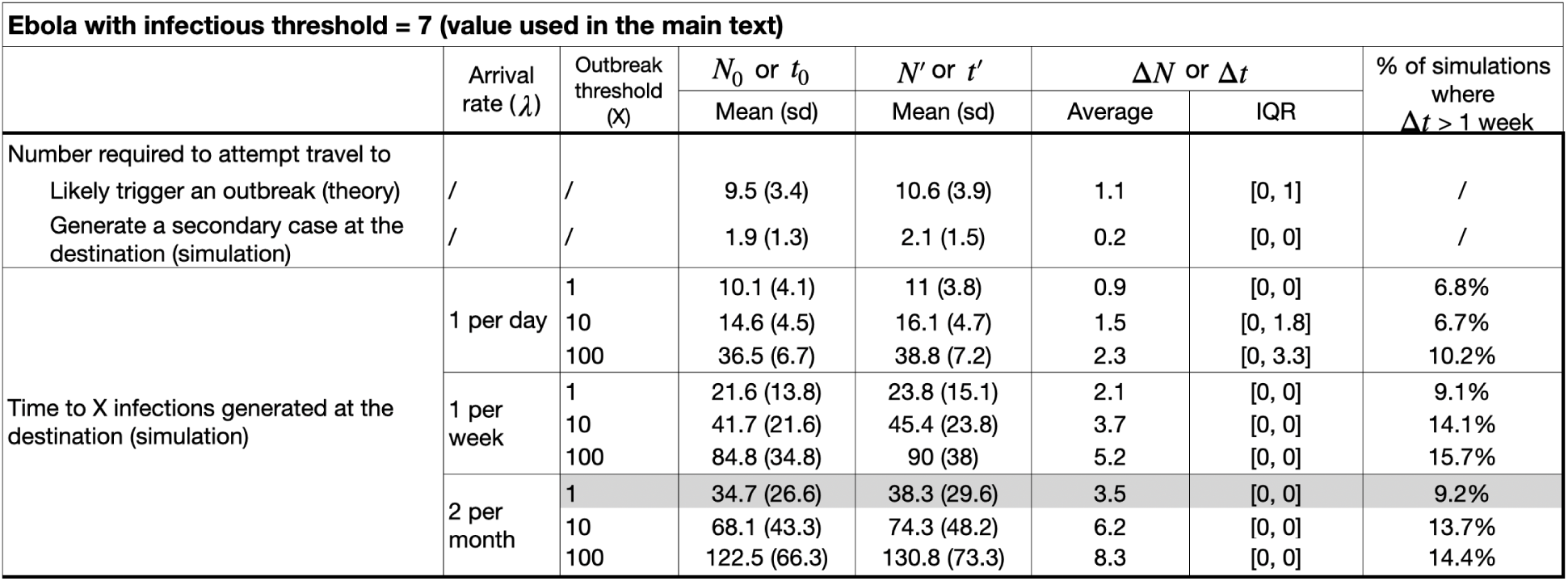
Screening effectiveness for Ebola. The number of infected travelers required to attempt travel to likely generate an outbreak with and without screening (*N^′^* and *N*_0_, respectively), and the time to *X* infections generated at the destination for a range of *X* with and without screening (*t^′^* and *t*_0_, respectively), and screening effectiveness Δ*N* = *N^′^ − N*_0_ and Δ*t* = *t^′^ − t*_0_ for a range of scenarios. The gray row is the plausible example reported in the Main Text.

**Figure S5:**
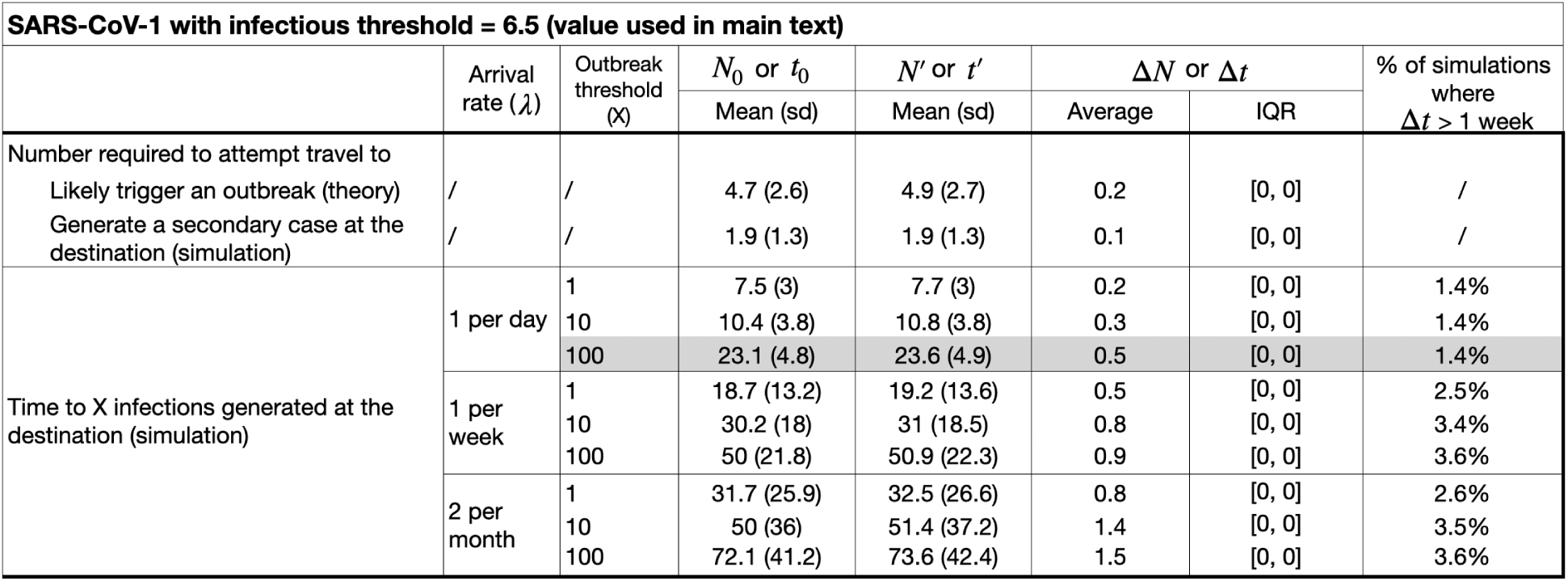
Screening effectiveness for SARS-CoV-1. The number of infected travelers required to attempt travel to likely generate an outbreak with and without screening (*N^′^* and *N*_0_, respectively), and the time to *X* infections generated at the destination for a range of *X* with and without screening (*t^′^* and *t*_0_, respectively), and screening effectiveness Δ*N* = *N^′^ − N*_0_ and Δ*t* = *t^′^ − t*_0_ for a range of scenarios. The gray row is the plausible example reported in the Main Text.

**Figure S6:**
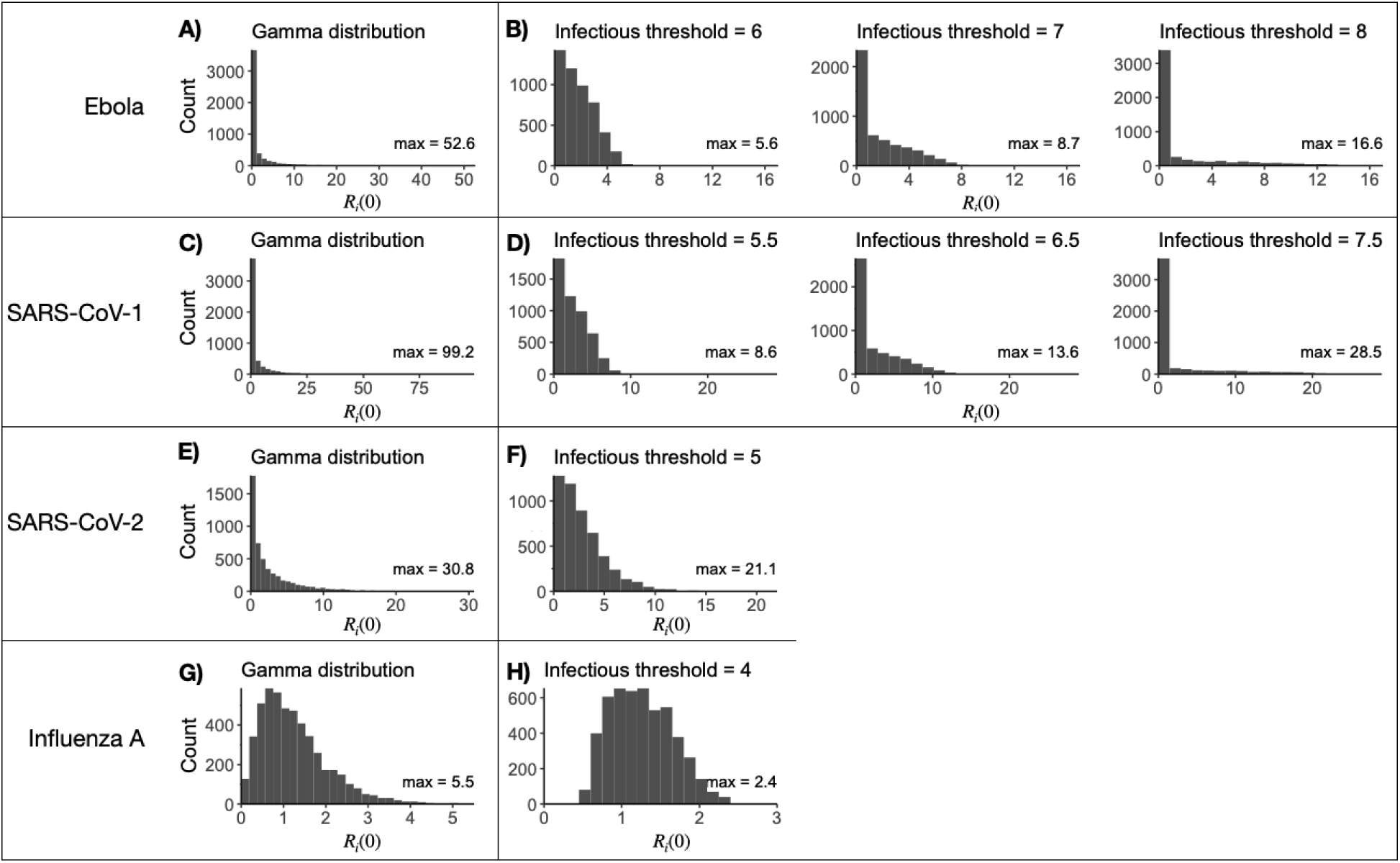
Different approaches to calculate the individual reproductive number *R_i_*(0) result in important differences in the population-level distributions of *R_i_*(0). (A, C, E, G) Distribution of the individual reproductive number *R_i_*(0) sampled from a gamma distribution with mean *R*_0_ and dispersion parameter *k*. (B, D, F, H) Distribution of *R_i_*(0) from the within-host viral kinetics model with various infectious thresholds.

**Figure S7:**
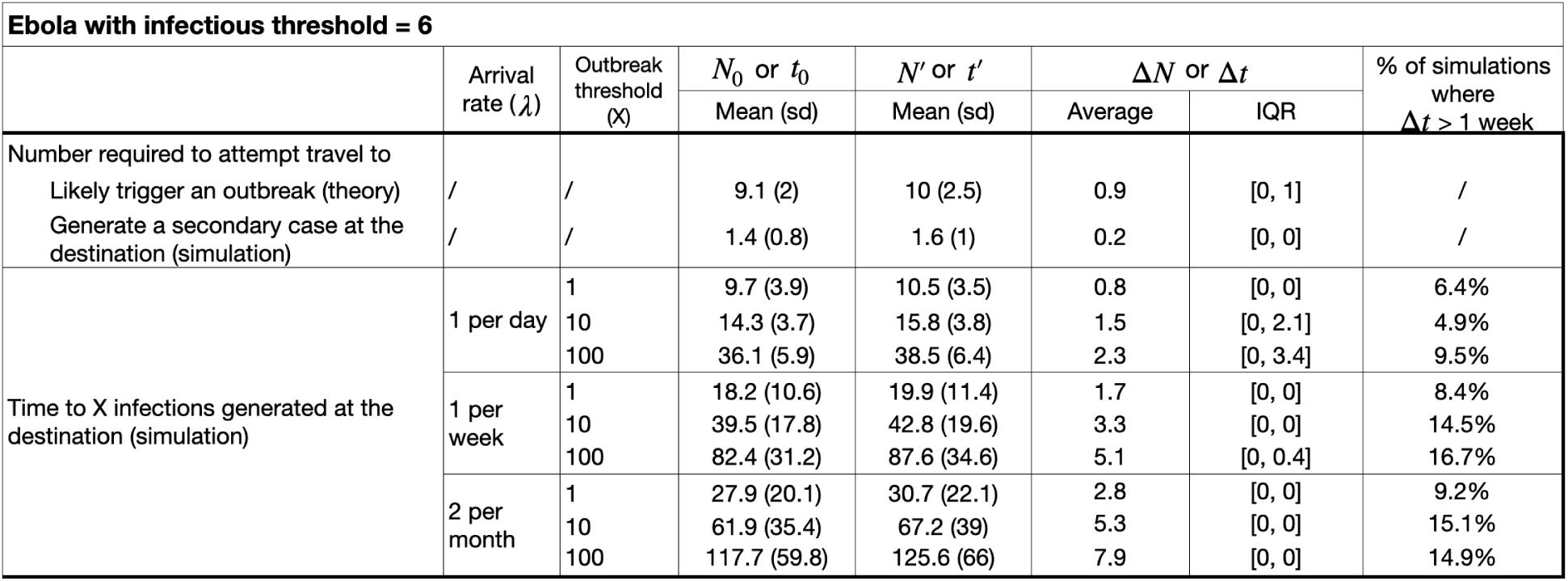
Screening effectiveness for Ebola with a lower infectious threshold. The number of infected travelers required to attempt travel to likely generate an outbreak with and without screening (*N^′^* and *N*_0_, respectively), and the time to *X* infections generated at the destination for a range of *X* with and without screening (*t^′^* and *t*_0_, respectively), and screening effectiveness Δ*N* = *N^′^ − N*_0_ and Δ*t* = *t^′^ − t*_0_ for a range of scenarios. The infectious threshold is 6 log10 copies RNA/mL. An infectious threshold of 7 log10 copies RNA/mL was used in the Main Text.

**Figure S8:**
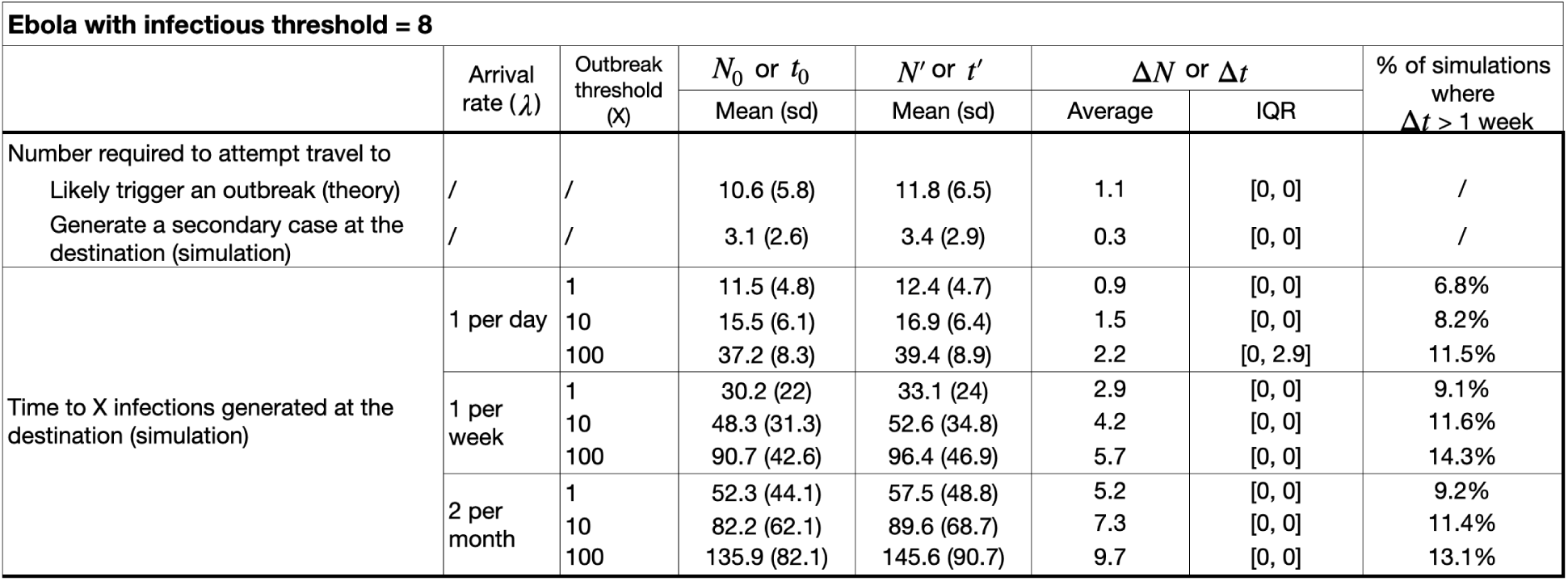
Screening effectiveness for Ebola with a higher infectious threshold. The number of infected travelers required to attempt travel to likely generate an outbreak with and without screening (*N^′^* and *N*_0_, respectively), and the time to *X* infections generated at the destination for a range of *X* with and without screening (*t^′^* and *t*_0_, respectively), and screening effectiveness Δ*N* = *N^′^ − N*_0_ and Δ*t* = *t^′^ − t*_0_ for a range of scenarios. The infectious threshold is 8 log10 copies RNA/mL. An infectious threshold of 7 log10 copies RNA/mL was used in the Main Text.

**Figure S9:**
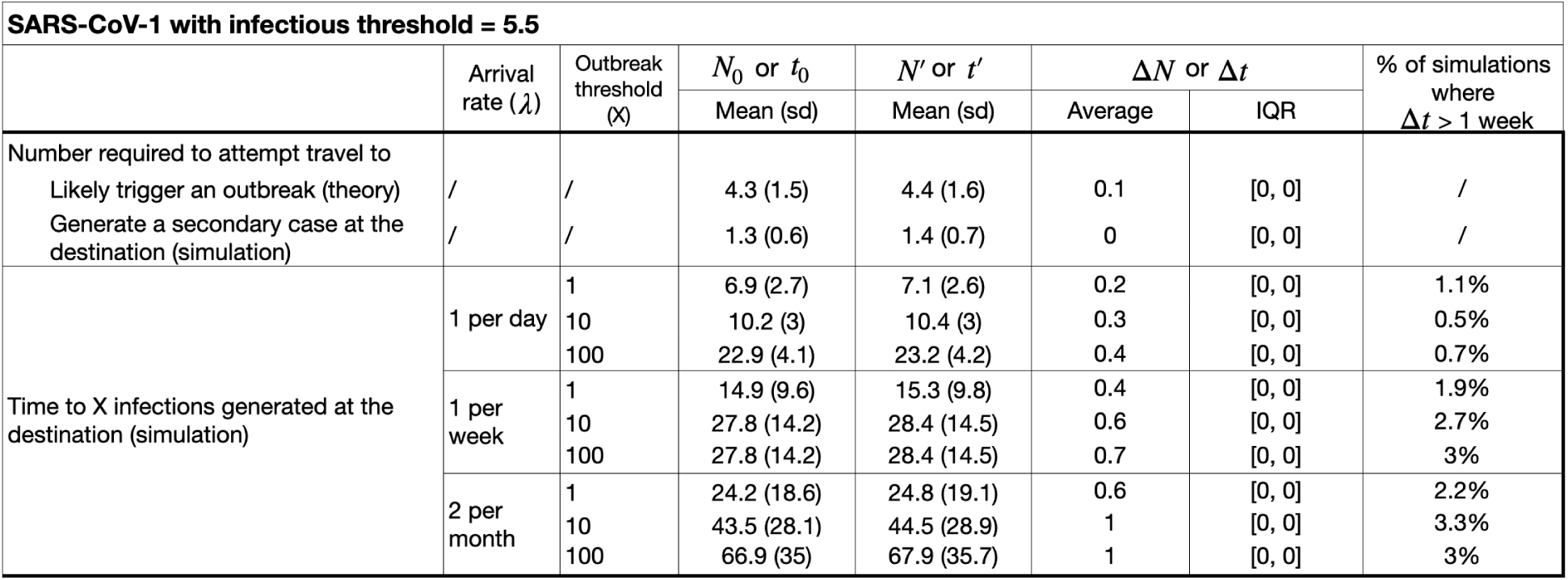
Screening effectiveness for SARS-CoV-1 with a lower infectious threshold. The number of infected travelers required to attempt travel to likely generate an outbreak with and without screening (*N^′^* and *N*_0_, respectively), and the time to *X* infections generated at the destination for a range of *X* with and without screening (*t^′^* and *t*_0_, respectively), and screening effectiveness Δ*N* = *N^′^ − N*_0_ and Δ*t* = *t^′^ − t*_0_ for a range of scenarios. The infectious threshold is 5.5 log10 copies RNA/mL. An infectious threshold of 6.5 log10 copies RNA/mL was used in the Main Text.

**Figure S10:**
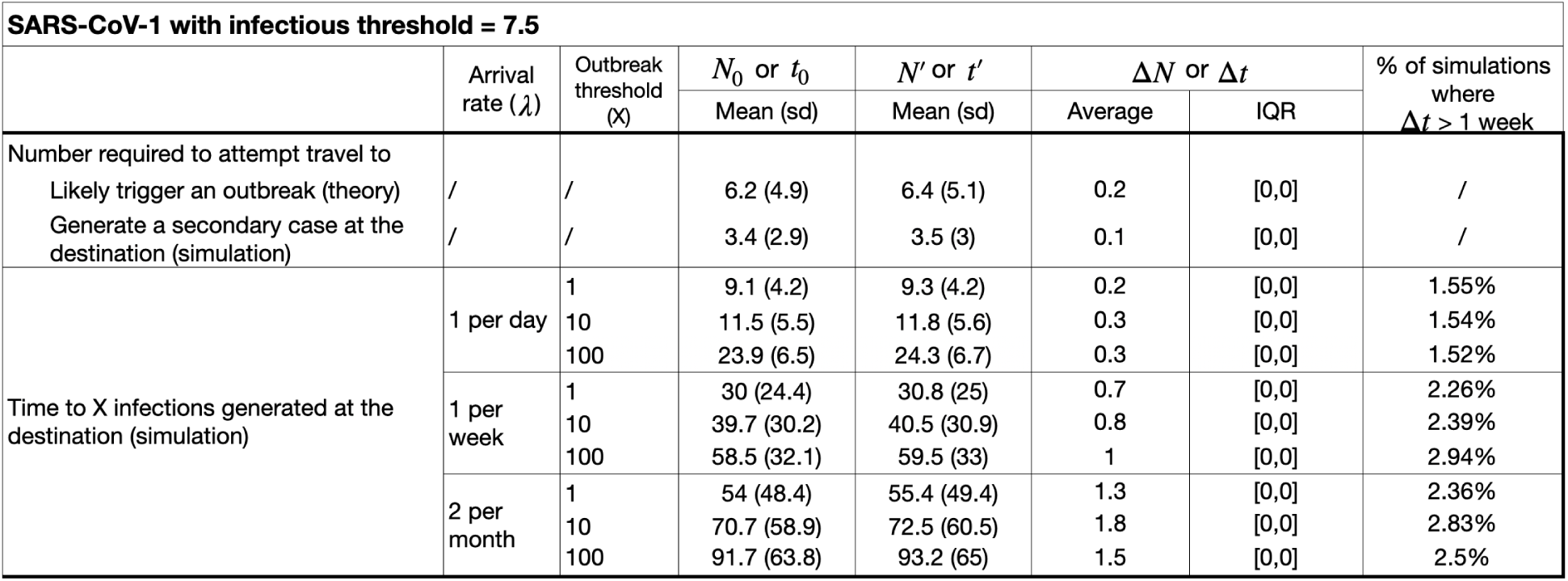
Screening effectiveness for SARS-CoV-1 with a higher infectious threshold. The number of infected travelers required to attempt travel to likely generate an outbreak with and without screening (*N^′^* and *N*_0_, respectively), and the time to *X* infections generated at the destination for a range of *X* with and without screening (*t^′^* and *t*_0_, respectively), and screening effectiveness Δ*N* = *N^′^ − N*_0_ and Δ*t* = *t^′^ − t*_0_ for a range of scenarios. The infectious threshold is 7.5 log10 copies RNA/mL. An infectious threshold of 6.5 log10 copies RNA/mL was used in the Main Text.

**Figure S11:**
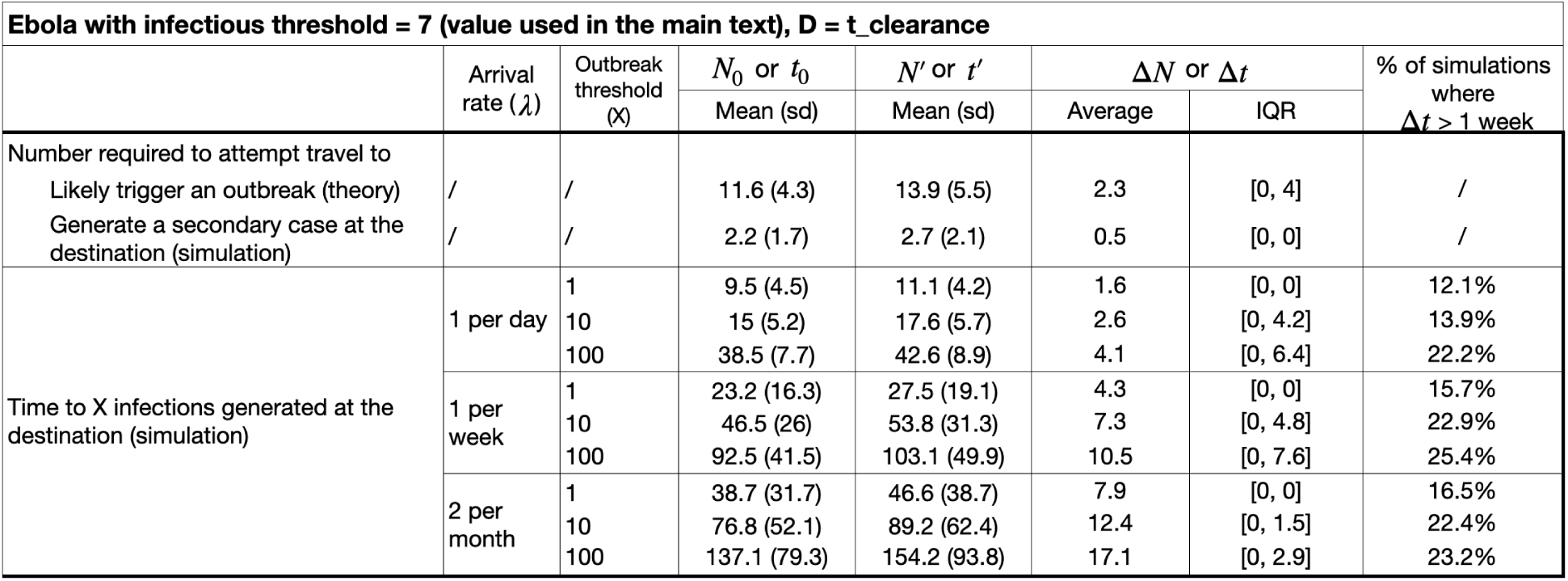
Screening effectiveness Δ*N* and Δ*t* for Ebola assuming people travel until viral clearance. The number of infected travelers required to attempt travel to likely generate an outbreak with and without screening (*N^′^* and *N*_0_, respectively), and the time to *X* infections generated at the destination for a range of *X* with and without screening (*t^′^* and *t*_0_, respectively), and screening effectiveness Δ*N* = *N^′^ − N*_0_ and Δ*t* = *t^′^ − t*_0_ for a range of scenarios. In the Main Text, we assumed people traveled up until the time they hospitalized.

**Figure S12:**
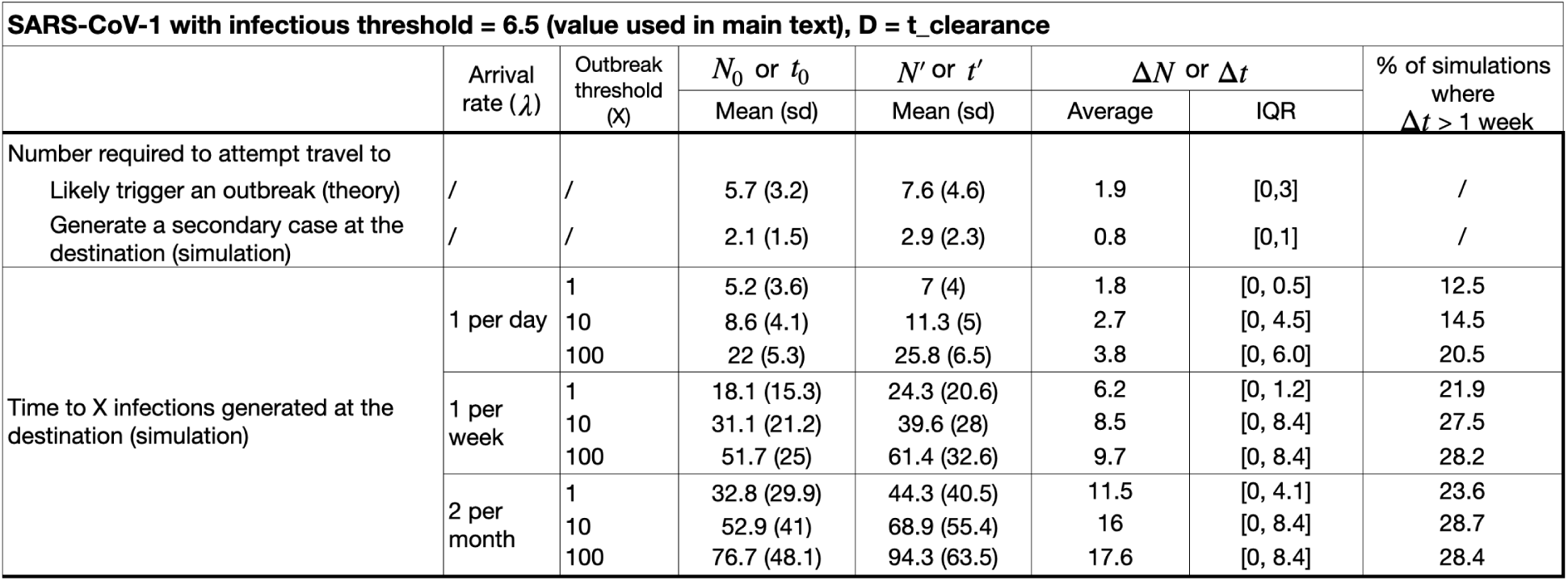
Screening effectiveness Δ*N* and Δ*t* for SARS-CoV-1 assuming people travel until viral clearance. The number of infected travelers required to attempt travel to likely generate an outbreak with and without screening (*N^′^* and *N*_0_, respectively), and the time to *X* infections generated at the destination for a range of *X* with and without screening (*t^′^* and *t*_0_, respectively), and screening effectiveness Δ*N* = *N^′^ − N*_0_ and Δ*t* = *t^′^ − t*_0_ for a range of scenarios. In the Main Text, we assumed people traveled up until the time they hospitalized.

**Figure S13:**
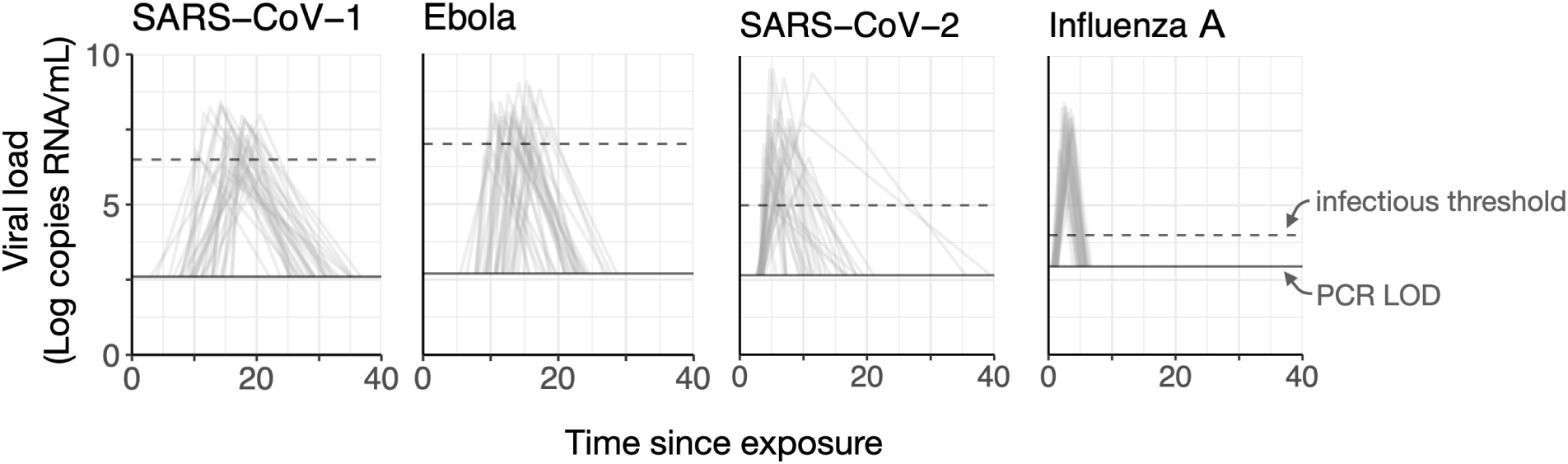
Simulated viral load trajectories for SARS-CoV-1, Ebola, SARS-CoV-2 and influenza. **A.** 100 stochastically drawn viral load trajectories for SARS-CoV-1, Ebola, SARS-CoV-2, and influenza A, using the control points and parameter values in Table S2.

